# Predicting Adverse Drug Effects: A Heterogeneous Graph Convolution Network with a Multi-layer Perceptron Approach

**DOI:** 10.1101/2022.03.22.22272749

**Authors:** Y.-H. Chen, Y.-T. Shih, C.-S. Chien, C.-S. Tsai

**Affiliations:** Dept. of Nephrology, Taichung Tzu Chi Hospital, Taichung, Taiwan; School of Medicine, Tzu Chi University, Hualien, Taiwan; Dept. of Applied Mathematics, National Chung Hsing University, Taichung, Taiwan; Dept. of Management of Information Systems, National Chung Hsing University, Taichung, Taiwan

## Abstract

The GCNMLP is implemented on three different datasets of side effects, namely, the SIDER, OFFSIDERS, and FAERS. Our results show that the performance of the GCNMLP on these three datasets is superior to the non-negative matrix factorization method (NMF) and some well-known machine learning methods with respect to various evaluation scales. Moreover, new side effects of drugs can be obtained using the GCNMLP.

**Author summary:** The GCNMLP enables us to get better drug side effect prediction, which improves personalized medicine prescriptions.

## Introduction

Adverse drug reaction (ADR) refers to the unexpected, harmful, or uncomfortable reactions of a drug intended to treat disease [1]. The U.S. Food and Drug Administration often removes prescription drugs from the market because of significant side effects [2], which result in enormous health and economic loss [3]. Traditionally, researchers document the side effects of drugs through cell or animal studies [4, 5], as the biochemical interrelationship between the expression and phenotypes of intracellular proteins is used to predict drug side effects at the laboratory development stage. However, these methods are often expensive and resource intensive. Furthermore, many serious side effects are not revealed until much later, highlighting the inadequacy of in vitro and in vivo studies.

Many studies in recent years have demonstrated the ability of machine learning algorithms to predict potential drug side effects [6, 7] by integrating different datasets [8, 9]. Established algorithms have used existing molecular biology databases such as gene expression and drug chemical properties to predict drug performance and possible side effects [10, 11], and the others have exploited internet search-keywords to infer potential side effects for monitoring drug safety [12]. In addition, natural language processing methodologies have also been applied to extract insights from scientific literatures, electronic medical records, and protein functional databases [13, 14]. Nevertheless, predicting side effects of drugs is still challenging, as drugs may affect multiple proteins, interfering subsequent protein networks [15].

Recently, link prediction methods with a matrix factorization approach have been used to predict drug side effects. Similar to matrix completion, each cell of the matrix represents whether a relationship may exist or not [16][p. 437–452]. For example, the matrix low-rank decomposition method based on singular value decomposition (SVD) and non-negative matrix factorization (NMF) are commonly used to solve the problems formulated as link prediction [17, 18]. Tensor factorization, like matrix factorization, can also be used to handle datasets with more than three dimensions to solve the link prediction [19]. The NMF has an advantage over the Bayesian networks since some prior information is required for the latter.

Link prediction can also be solved using the node2vec algorithm. As similar nodes tend to aggregate together, the nodes can be encoded with a biased random walk to represent the features of neighboring nodes. This can further be exploited to predict link existence [20]. This method has also been adopted for drug repositioning from known drug-disease relationships using a heterogeneous network with collaborative filtering [21]. Another study applied a heat diffusion kernel method to predict the relationships among genes, protein function, and disease, achieving an area under receiver operating characteristic curve (AUROC) value of 92.3% and reducing the error rate by 52.8% [22]. These studies demonstrate that exploring drugs with similar protein binding properties may pave a new direction for verifying drug reliability and effectiveness.

Network analyses that use link prediction include Random walk [23] and PageRank [24]. Random walk was applied to predict drug responses in cell lines which classified sensitive and drug resistant cell lines with 85% accuracy [25]. Similarly, graph neural networks have been used in disease prediction and classification [26], in biological information problems [27], in social recommendation systems [28] as well as in knowledge graph applications [29]. Convolutional neural network (CNN) is another widely implemented method for extracting spatial patterns in data or images [30].Graph convolution network (GCN) is a similar method, but it operates on graphs [31].

During the past years, machine learning and deep neural networks have been exploited to study the prediction of drug side effects. For instance, Zitnik *et al*. [32] proposed a model called Decagon for dealing with the prediction of polypharmacy interactions. However, it could not predict the side effects of a single drug. Muñoz *et al*. [33] exploited knowledge graphs and multi-label learning models to reveal potential drug adverse reactions with AUPR=0.429 and AUROC=0.886. Their approach is flexible and can be tuned for specific requirements. Mohsen *et al*. [34] applied deep neural networks to predict ADR in the datasets TG-GATEs and FAERS incorporating the the gene expression profiles. Dey *et al*. [35] proposed some machine learning models including a deep learning one which could predict ADRs and identified the molecular substructures associated with the ADRs without defining the substructures *a-priori* in advance. In addition, Guney [36] implemented several machine learning methods such as logistic regression, *k*-nearest neighbor classier, support vector machine, random forest, and gradient boosting classifier on the datasets Drugbank [37], PubChem [38], and SIDER [39]. The mean value of AUROC is 0.841, and that of the AUPR is 0.837 on these datasets. Furthermore, Galeano and Paccanaro [40] used the collaborative filtering model to predict drug side effects on the dataset SIDER with 1,525 marketed drugs and 2,050 side effects, and obtained AUPR=0.342.

In this paper we apply a heterogeneous GCN combined with a multi-layer perceptron (MLP), which is denoted by GCNMLP, to explore potential side effects of drugs. Note that any dataset presented as a network structure can be embedded via GCN for downstream machine learning application. The aggregation of nodes and edges characteristics generates representation learning from the graph, node classification, link prediction, and other tasks [26]. The GCN has been applied to social network research [28], biomedicine networks [41] and knowledge graphs [42][p. 593–607]. By inferring the relationship among similar drugs, the GCNMLP shortens the time consumption in uncovering the side effects unobserved in routine drug prescriptions. Our results predict drug side effects with AUPR = 0.941, which is superior to other well-known methods used in the literatures [25, 43]. In addition, new side effects which were not found in original dataset can be obtained using the GCNMLP.

### Datasets

The datasets in this study were downloaded from open resources, where the drug information was obtained from DrugBank Online [44](https://go.drugbank.com/), and the side effects information from three Adverse Drug Event (ADE) on three databases: the Side Effect Resource (SIDER) [45](http://sideeffects.embl.de/), OFFSIDES [46] (http://www.pharmgkb.org/downloads.jsp), and the United States Food and Drug Administration (FDA) Adverse Event Reporting System (FAERS) (https://open.fda.gov/data/faers/). The datasets of side effects (refer to github.com/yishingene/gcnmlp) contains four columns: ‘drugbank_id’ is the identification number of the database from the University of Alberta, ‘drugbank_name’ is the drug name, ‘umls cui from meddra’ is the coded number of the Unified Medical Language System, and ‘side_effect_name’ is the reported side effects. Concerning the dataset SIDER there are 4245 drugs, 17671 side effects as well as 3,766,382 drug-side effect associations for positive links. Similar information of the datasets OFFSIDES and FAERS can be found in Table 1. The remaining 71,247,013 possible association between drug and side effects are negative links. There are three columns in the file ‘semantic_similarity_side_effects_drugs’. The first and the second columns are the pairs of drug names. The third column is the similarity score of drugs from the first and the second columns. Similarity scores between the drugs in the dataset are the cosine similarity derived from the word2vec package combined with a model pre-trained on PubMed, PMC, and Wikipedia texts by the work of Mohsen *et al*. [34]. The word representations are derived from the large corpus of biomedical and general-domain texts, including PubMed abstracts (nearly 23 million abstracts) and PMC articles (about 700000 full texts), plus approximately four million English Wikipedia articles. The process of combining these datasets is illustrated in Fig 1.

**Table 1.** Statistics of adverse drug event association pairs in three datasets

| Dataset | No of drugs | No of ADE | No of pairs |
| --- | --- | --- | --- |
| SIDER | 1020 | 5599 | 133,750 |
| OFFSIDES | 2738 | 14544 | 3,206,558 |
| FAERS | 4245 | 17671 | 3,766,382 |

**Fig 1.**
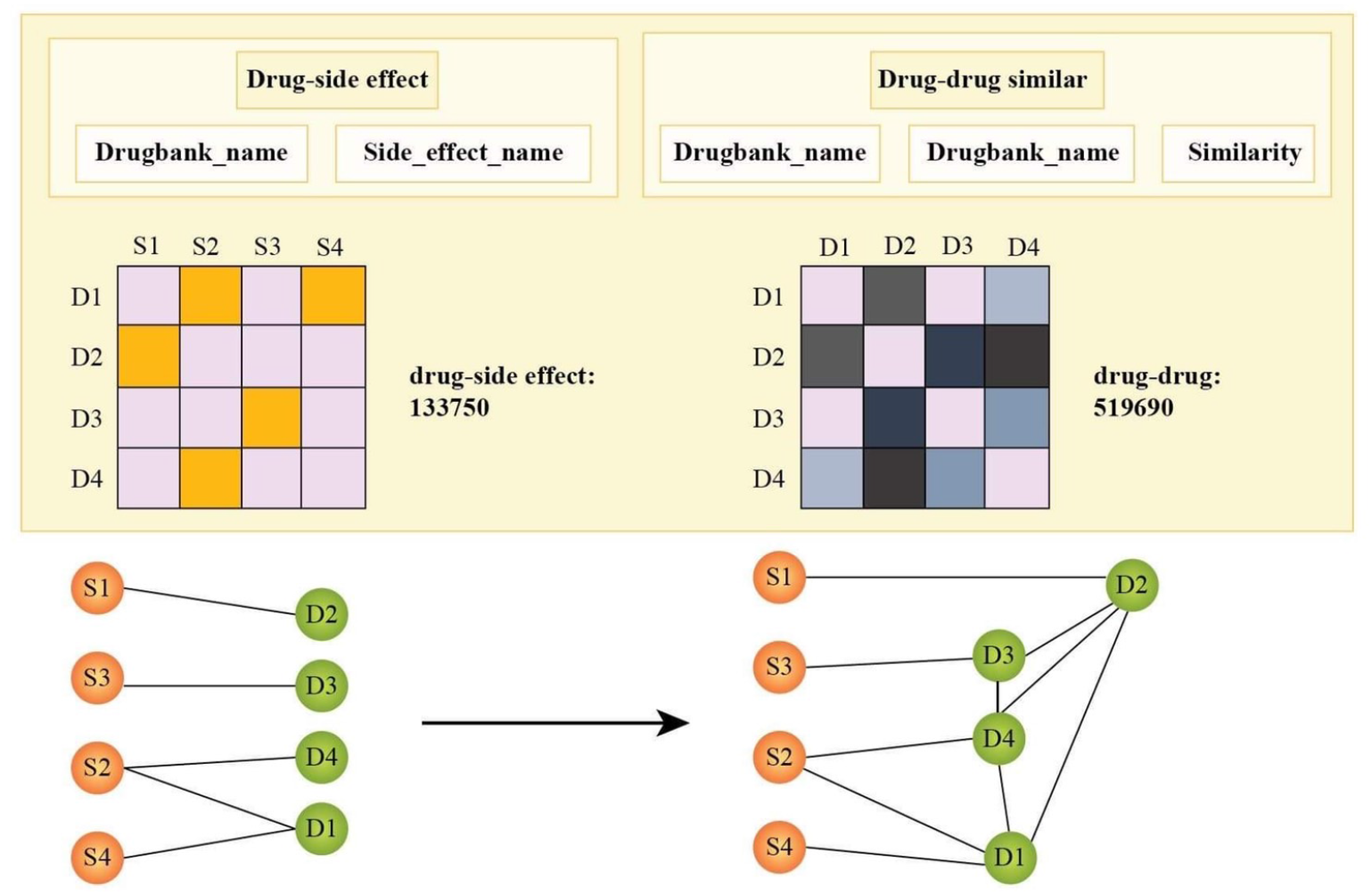
The illustration of the combination of datasets.

### NMF-based and link prediction methods

For comparison we briefly review some well-known algorithms concerning link prediction.

#### i. Non-negative matrix factorization (NMF)

For convenience we define the set 𝕊 ≡ ℝ^+^ ∪{0}, where ℝ^+^ is the set of positive real numbers. Let the relationship between drugs and side effects be described by a bipartite matrix **V** ∈ 𝕊 ^*m*×*n*^. We factor **V** as

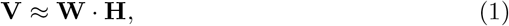

where the matrices **W** = (*w*_*ij*_) ∈ 𝕊 ^*m*×*k*^ and **H** = (*h*_*jl*_) ∈ 𝕊 ^*k*×*n*^ are low-rank approximations with dimension *k* ≪ min {*m, n*}, and the recovered elements *w*_*ij*_ and *h*_*jl*_ in the matrices are the potential links. Note that low-rank matrix factorization could reveal potential links between drugs and side effects [43].

#### ii. Non-negative matrix factorization with heat diffusion (NFMHD)

We apply the heat diffusion equation to the bipartite matrix concerning drug side effects with drug-drug similarity matrix. Consider an undirected network graph *G* = (𝒱, ℰ, 𝒫), where 𝒱 = {*v*_1_, ℰ = *v*_2_, …, *v*_*n*_} is the set of nodes, 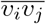: for any two connecting nodes *v*_*i*_ and *v*_*j*_, 1 ≤ *i, j* ≤ *n, i* ≠ *j*, } the set of all edges, and 𝒫 = {*ω*_*ij*_} is the probability space which is the collection of the link probability *ω*_*ij*_ for the existence of all edges, 0 ≤ *ω*_*ij*_ ≤ 1. Let **f** (*t*) be the link vector connecting the nodes *v*_*i*_ and *v*_*j*_ at time *t*, and **f** (0) an initial value at time zero. The discrete approximation for the heat diffusion flow [43, 48] is expressed as

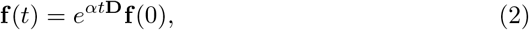

where *α* is the thermal conductivity coefficient, and **D** = (*d*_*ij*_) is the diffusion matrix for the node *v*_*j*_ of the undirected graph, which is given by

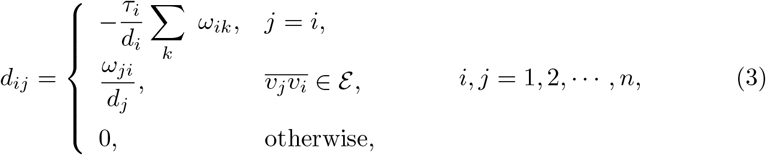

where the degrees *d*_*j*_ ≥ 1, and *τ*_*j*_ is the flag index to the out-line, 0 ≤ *τ*_*j*_ ≤ 1. Note that the diffusion matrix is derived from the drug-drug similarity matrix.

### Heuristic network link-prediction methods

#### iii. Adamic Adar (AA) [49]

The Adamic Adar computes the probability of two linking nodes *v*_*i*_ and *v*_*j*_ by finding the common neighbors of these two nodes, and calculating the sum of the inverse logarithmic degree of the common neighbors by

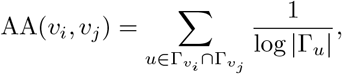

where Γ_*u*_ is the set of neighbors of node *u*.

#### iv. Resource allocation (RA)

The resource allocation algorithm [50] is designed for predicting the links between two nodes *v*_*i*_ and *v*_*j*_ by measuring the sum of the inverse degree of the common neighbors, and is given by

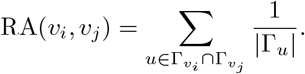

#### v. Katz

The Katz algorithm [51] is a path-based method. The probability of existing links between two nodes *v*_*i*_ and *v*_*j*_ depends on the number of paths between the two nodes, and is given by

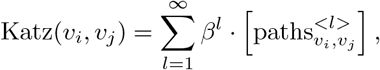

where *β* represents the attenuation factor, 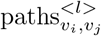 is the degree of connection between the nodes *v*_*i*_ and *v*_*j*_ through some walk of length *l*. Hence the Katz’s path is exponentially controlled by the path length, and more weight is required for shorter path.

#### vi. Personalized PageRank (PPR)

The Personalized PageRank PPR(*v*_*i*_, *v*_*j*_) [52, 53] calculates either the likelihood of a random walk from the node *v*_*j*_ to the node *v*_*i*_ with a probability *ω* or a random neighbor with a probability 1 − *ω*.

### The GCNMLP

In this section we describe an end-to-end GCNMLP to predict potential relationships between drugs and side effects. Let 𝒢 = (𝒱, ℰ, 𝒫) be a bipartite graph which represents the network between drugs and side effects, where the sets 𝒱, ℰ and 𝒫 are defined as in Section 2. A homogeneous graph which integrates the similarities between drugs and drugs is attributed to the bipartite network. Let ℳ ⊂ ℝ^*m*×*n*^ be a real matrix which represents a bipartite graph of drugs and side effects, where *m* is the number of drug effects and *n* is the number of side effects. Let *p*, 0 ≤ *p* ≤ *m*, be a row index which denotes the drug effect index number, and *q*, 0 ≤ *q* ≤ *n*, a column index representing the side effect index number. The relationship between drugs and side effects is given by *ω*_*p,q*_ ∈ 𝒫, 0 ≤ *ω*_*p,q*_ ≤ 1. In addition, *ω*_*p,q*_ = 0 means that there is no connection, and *ω*_*p,q*_ = 1 stands for the connection between drugs and side effects. Otherwise *ω*_*p,q*_ denotes the unknown predicted value.

The spatial-based method of GCN is similar to that of the convolutional neural network (CNN), where the representations from the neighboring nodes of a given node *ν* are aggregated and updated. Moreover, it also outputs the representations of the graph required [54]. The GCNMLP is formulated as

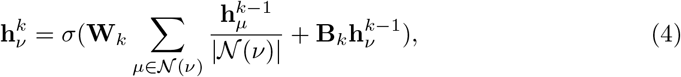

where 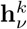 represents the feature of the link between the node *ν* ∈ 𝒱 and the set of neighbors 𝒩(*ν*) with weighting matrices **W**_*k*_ and **B**_*k*_, and *σ* is a differentiable aggregator function in the *k*-th aggregation step. Equation (4) means that the node *ν* aggregates representations from each node in the neighborhood *𝒩*(*ν*) on the previous (*k* − 1)-th aggregation step, which is given by 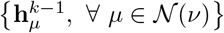. When *k* = 0 the representations are just the initial input of the link. Equation (4) can further be generalized as

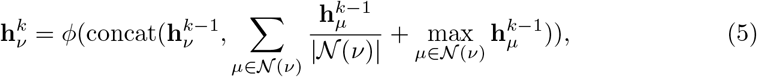

where *ϕ*(·) is either a function of a multi-layer perceptron, or dot product of neighboring nodes, or other self-defined functions to represent the link through node features, and ‘concat’ is the concatenate function where the input features are concatenated. The link probability between two adjacent nodes is predicted as a probability from the node’s features. Let 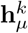 and 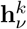 be the features of two adjacent nodes *μ* and *ν*, respectively. We define Φ as a function of the dot product of 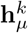 and 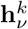 or the multi-layer perceptron. The score of the link between the nodes *μ* and *ν* is defined as

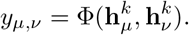

The architecture of the GCNMLP is illustrated in Fig 2.

**Fig 2.**
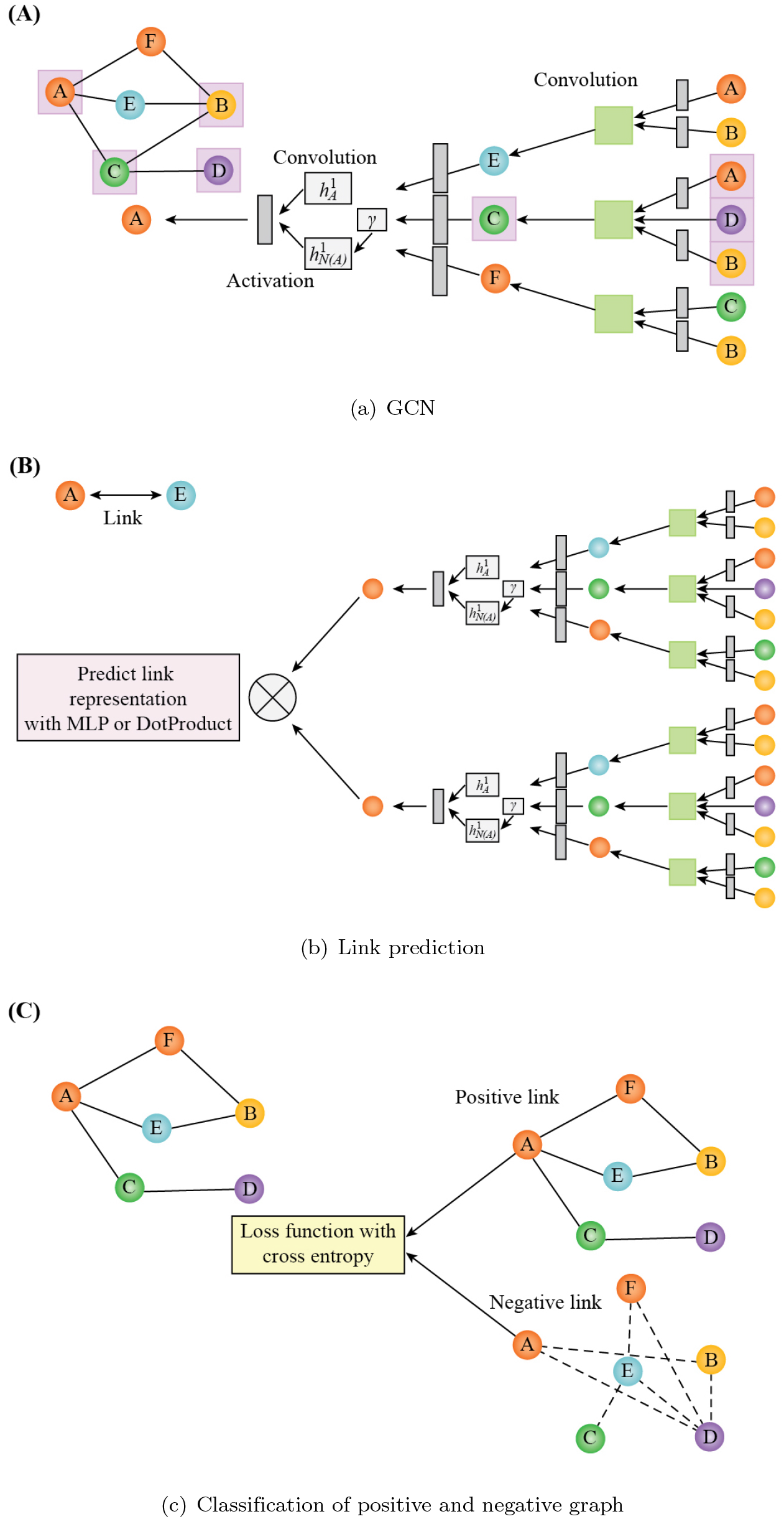
The architecture of the GCNMLP.

### Experimental Results

In this section, we implemented the GCNMLP on the three datasets SIDER, OFFSIDES, and FAERS for link prediction. The GCN was applied to assess the predictability of drug side effects, and the MLP was exploited to classify the positive and negative graphs. In addition, the dataset was shuffled before being split into 90% for training and 10% for testing 10-fold cross-validation procedure to test our predictive model. We used the Precision, Recall, F1 score, AUROC, and AUPR as evaluation scales. All experiments were run 10 times including the mean results and the standard deviations.

#### Comparing different dropout rates in the GCNMLP

The dropout rate is important if we wish to improve the performance of deep learning models. Different dropout rates with 0%, 10%, 30%, 50% were used in our model. We observe from Table 2 that a trend of deteriorating performance occurred on all evaluation scales when the dropout rate was increasing.

**Table 2.** Comparing different dropout rates in the GCNMLP.

| Dropout | Precision | Recall | F1 score | AUROC | AUPR |
| --- | --- | --- | --- | --- | --- |
| 0% | 0.865±0.003 | 0.865±0.003 | 0.865±0.003 | 0.939±0.002 | 0.941±0.002 |
| 10% | 0.862±0.003 | 0.858±0.003 | 0.858±0.003 | 0.934±0.002 | 0.932±0.003 |
| 30% | 0.857±0.003 | 0.853±0.003 | 0.853±0.003 | 0.926±0.002 | 0.926±0.002 |
| 50% | 0.848±0.003 | 0.843±0.003 | 0.842±0.003 | 0.917±0.002 | 0.918±0.003 |

#### Comparing different percentage of the training set in the GCNMLP

Various data sizes of the training set were considered in our experiments. From Table 3 we find that when the size of the training set was decreased from 90% to 80%, the performance on the GCNMLP on all the evaluation scales deteriorates when using the 10-fold cross-validation. When the size of the training set was decreased from 80% to 50%, the performance of these two training sets almost makes no difference on all evaluation scales except the AUPR. However, the deviation of the latter is smaller than that of the former.

**Table 3.** Comparing different percentage of the training set in the GCNMLP.

| Training | Precision | Recall | F1 score | AUROC | AUPR |
| --- | --- | --- | --- | --- | --- |
| 90% | 0.865±0.003 | 0.865±0.003 | 0.865±0.003 | 0.939±0.002 | 0.941±0.002 |
| 80% | 0.864±0.004 | 0.860±0.004 | 0.860±0.004 | 0.937±0.002 | 0.937±0.004 |
| 50% | 0.864±0.000 | 0.860±0.000 | 0.860±0.000 | 0.937±0.000 | 0.938±0.0000 |

#### Comparing different hop numbers in the GCNMLP

We implemented the GCNMLP with different hop numbers. Table 4 shows that the GCNMLP performs best with the hop number being 3. When the hop number *n* = 4 or 5, the performance of the GCNMLP are almost the same on all the evaluation scales except the AUPR. But the deviation of the latter is greater than that of the former.

**Table 4.** Comparing different hop numbers in the GCNMLP.

| hop # | Precision | Recall | F1 score | AUROC | AUPR |
| --- | --- | --- | --- | --- | --- |
| $n = 2$ | $0.863 \pm 0.003$ | $0.859 \pm 0.003$ | $0.859 \pm 0.003$ | $0.935 \pm 0.003$ | $0.935 \pm 0.003$ |
| $n = 3$ | $0.865 \pm 0.003$ | $0.865 \pm 0.003$ | $0.865 \pm 0.003$ | $0.939 \pm 0.002$ | $0.941 \pm 0.002$ |
| $n = 4$ | $0.864 \pm 0.002$ | $0.864 \pm 0.002$ | $0.864 \pm 0.002$ | $0.938 \pm 0.002$ | $0.939 \pm 0.003$ |
| $n = 5$ | $0.864 \pm 0.003$ | $0.864 \pm 0.003$ | $0.864 \pm 0.003$ | $0.938 \pm 0.003$ | $0.940 \pm 0.004$ |

#### Comparing the performance of various algorithms

We compared the performance of the GCNMLP to NMF,NMFHD, and some link-prediction methods. Table 5 shows that the GCNMLP performs best among these methods on Recall, F1 score, and AUPR. In addition, the non-negative matrix factorization methods are superior to the heuristic network link-prediction methods on these three items. On the other hand, the NMF outperforms the other methods on AUROC, and the NMFHD is superior to the other methods mentioned above on Precision. Fig 3 displays the AUPR score of various algorithms. Fig 4(a) shows that the NMF performs best on the receiver operating characteristic (ROC) curve among the other methods mentioned above. However, the GCNMLP is quite competitive compared to that of the NMF and NMFHD. Fig 4(b) displays that the GCNMLP is superior to all the other methods on the AUPR. From the results mentioned above it is obvious that the heuristic network link-prediction methods are inferior to the GCNMLP and the non-negative matrix factorization methods on all the items.

**Table 5.** Comparing the performance of various algorithms.

| Model | Precision | Recall | F1 score | AUROC | AUPR |
| --- | --- | --- | --- | --- | --- |
| GCNMLP | $0.865 \pm 0.003$ | **$0.865 \pm 0.003$** | **$0.865 \pm 0.003$** | $0.939 \pm 0.002$ | **$0.941 \pm 0.002$** |
| (Non-negative matrix factorization methods) |  |  |  |  |  |
| NMF | $0.906 \pm 0.003$ | $0.659 \pm 0.001$ | $0.726 \pm 0.002$ | **$0.946 \pm 0.001$** | $0.600 \pm 0.003$ |
| NMFHD | **$0.930 \pm 0.002$** | $0.623 \pm 0.002$ | $0.689 \pm 0.002$ | $0.943 \pm 0.001$ | $0.585 \pm 0.003$ |
| (Heuristic network link-prediction methods) |  |  |  |  |  |
| AA | $0.840 \pm 0.008$ | $0.525 \pm 0.003$ | $0.541 \pm 0.006$ | $0.909 \pm 0.001$ | $0.334 \pm 0.006$ |
| PA | $0.593 \pm 0.076$ | $0.501 \pm 0.001$ | $0.496 \pm 0.002$ | $0.916 \pm 0.001$ | $0.374 \pm 0.007$ |
| RA | $0.490 \pm 0.006$ | $0.500 \pm 0.000$ | $0.494 \pm 0.000$ | $0.909 \pm 0.001$ | $0.334 \pm 0.006$ |
| PPR | $0.836 \pm 0.007$ | $0.542 \pm 0.002$ | $0.570 \pm 0.003$ | $0.908 \pm 0.001$ | $0.351 \pm 0.004$ |
| Katz | $0.778 \pm 0.005$ | $0.587 \pm 0.004$ | $0.629 \pm 0.005$ | $0.909 \pm 0.002$ | $0.353 \pm 0.005$ |

**Fig 3.**
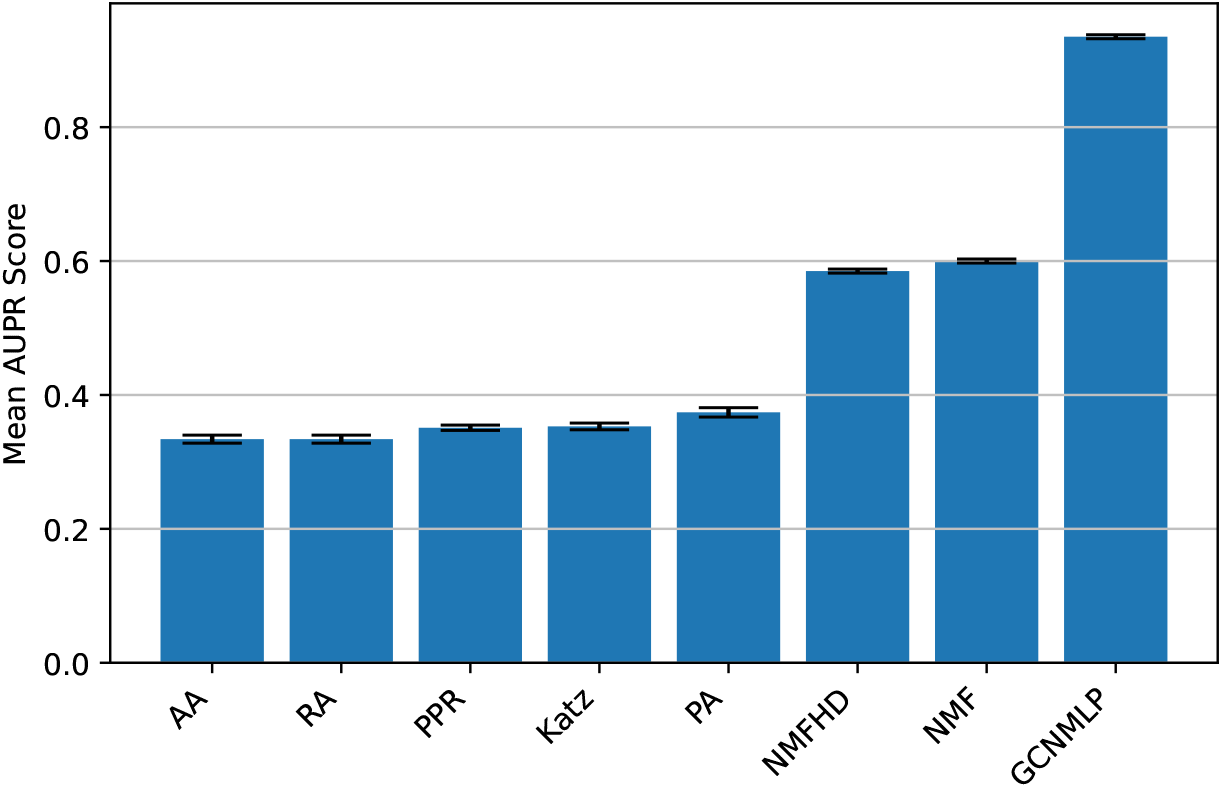
Comparing the performance of various algorithms on AUPR (score).

**Fig 4.**
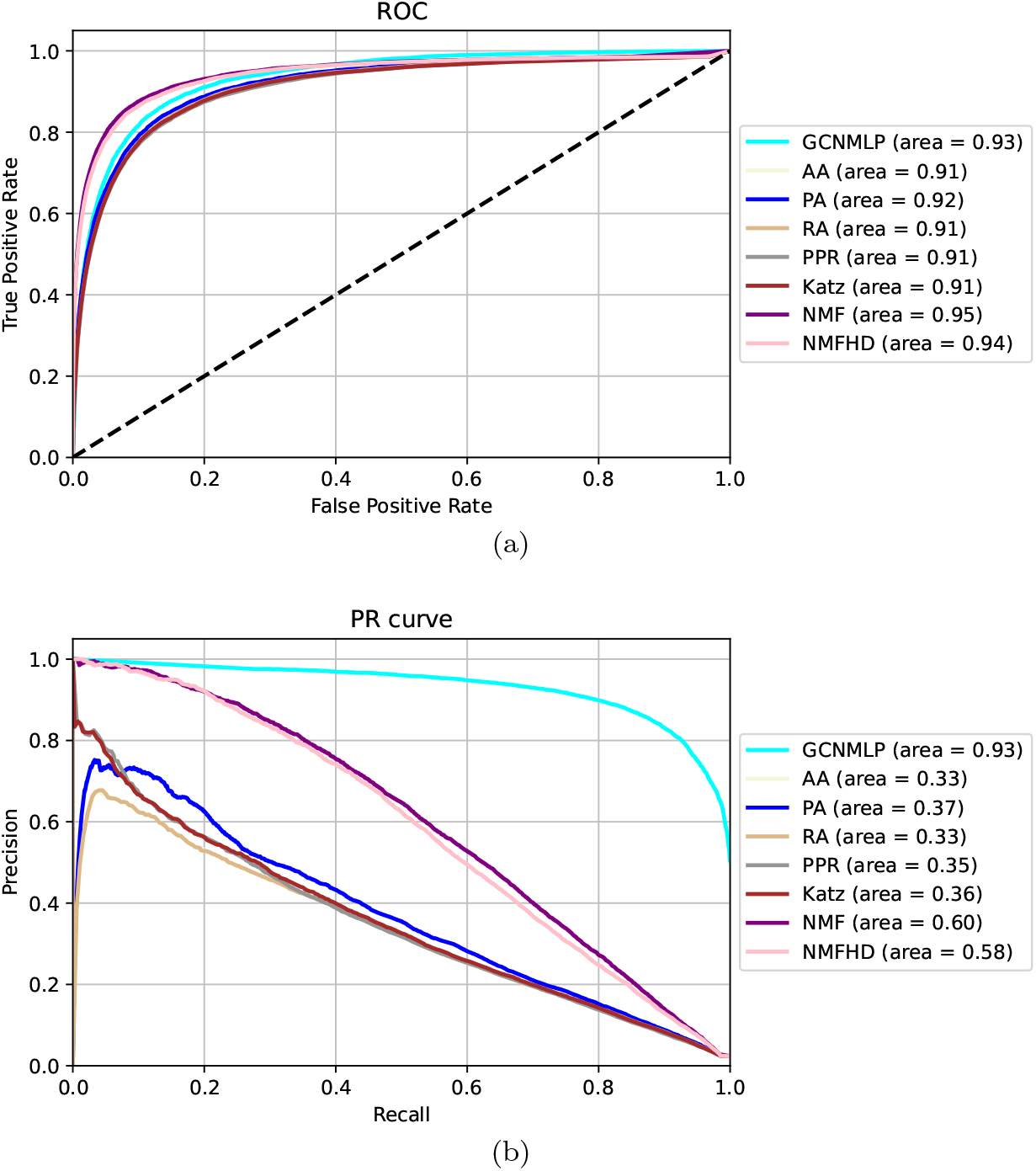
Comparing the performance of various algorithms on the ROC and PR curves.

#### Comparing the performance of the GCNMLP on the three datasets

Table 6 shows that the GCNMLP performs best with respect to all the evaluation scales on FAERS, and the results on OFFSIDES are better than their counterparts on SIDER. From Tables 6–7 we see that the performance of the GCNMLP on the three datasets with respect to AUROC and AUPR are superior to various machine learning methods such as collaborative filtering, logistic regression, *k*-nearest neighbor classifier, support vector machine, random forest, and gradient boosting classifier. In addition, the performance of the gradient boosting classifier is better than that of the other machine learning methods on AUROC and AUPR.

**Table 6.** Comparing the performance of the GCNMLP on the three datasets

| Dataset | AUROC | AUPR | Precision | Recall | F1 score |
| --- | --- | --- | --- | --- | --- |
| SIDER | $0.939 \pm 0.002$ | $0.941 \pm 0.002$ | $0.865 \pm 0.003$ | $0.865 \pm 0.003$ | $0.865 \pm 0.003$ |
| OFFSIDES | $0.964 \pm 0.009$ | $0.955 \pm 0.013$ | $0.902 \pm 0.011$ | $0.903 \pm 0.012$ | $0.902 \pm 0.012$ |
| FAERS | **$0.973 \pm 0.002$** | **$0.972 \pm 0.002$** | **$0.913 \pm 0.001$** | **$0.913 \pm 0.001$** | **$0.913 \pm 0.001$** |

**Table 7.** Comparing the performance of the GCNMLP with various machine learning methods for SIDER

| Model | AUROC | AUPR |
| --- | --- | --- |
| GCNMLP | **$0.939 \pm 0.002$** | **$0.941 \pm 0.002$** |
| collaborative filtering | $0.901 \pm 0.002$ | $0.609 \pm 0.003$ |
| logistic regression | $0.898 \pm 0.058$ | $0.836 \pm 0.126$ |
| k-nearest neighbor classifier | $0.882 \pm 0.090$ | $0.764 \pm 0.161$ |
| support vector machine | $0.898 \pm 0.048$ | $0.836 \pm 0.106$ |
| random forest | $0.889 \pm 0.069$ | $0.799 \pm 0.125$ |
| gradient boosting classifier | $0.906 \pm 0.043$ | $0.843 \pm 0.080$ |

Finally, we performed the t-test to find out if there was a significant difference in the 10 fold cross validation results among the GCNMLP and the NMF based methods and the graph-based link prediction methods using a significance level with *α* = 0.05. After the t-test was computed, we computed and compared to the significance level with *α* = 0.05. If the p-value was smaller than *α*, we rejected the null hypothesis. Table 8 shows the testing results of the p-values.

**Table 8.** Comparing the p-value of the t-test among the GCNMLP and various algorithms

| Model | Precision | Recall | F1 score | AUROC | AUPR |
| --- | --- | --- | --- | --- | --- |
| GCNMLP vs AA | 7.430e-08 | 5.703e-33 | 2.019e-29 | 1.433e-15 | 1.766e-33 |
| GCNMLP vs PA | 3.063e-09 | 3.129e-36 | 5.829e-35 | 2.094e-13 | 1.270e-32 |
| GCNMLP vs RA | 7.124e-31 | 8.030e-37 | 7.201e-37 | 1.423e-15 | 1.816e-33 |
| GCNMLP vs PPR | 7.042e-09 | 6.455e-35 | 4.056e-33 | 3.306e-16 | 4.561e-38 |
| GCNMLP vs Katz | 2.946e-20 | 8.275e-31 | 3.736e-29 | 1.113e-15 | 9.453e-36 |
| GCNMLP vs NMF | 3.459e-19 | 8.663e-31 | 2.311e-27 | 6.940e-10 | 6.256e-33 |
| GCNMLP vs NMFHD | 1.667e-22 | 2.187e-32 | 1.928e-29 | 1.852e-07 | 1.391e-33 |

## Discussion

Non-negative matrix factorization methods perform well on the evaluation scales Precision and AUROC. In particular, the NMFHD performs best on the scale Precision among all the methods implemented in this paper including the results of the GCNMLP on the three datasets. Next, machine learning methods are superior to link-predictions and non-negative matrix factorization methods on the scale AUPR. Conversely, link-prediction methods are superior to machine learning methods on the scale AUROC. See Tables 5 and 7 for details.

In our experiments we have inquired four drugs from the trained model, where the parameters used were acquired from the dataset SIDER after training. The probability scores of the ten highest-ranked side effects of these drugs were calculated and ranked. See Tables 9–10 for details. From Table 9 we see that the side effects diarrhea, dyspepsia, and musculoskeletal discomfort associated with the drug amlodipine which have been reported in [57], [58, 59], and [60], respectively, were detected in our experiment. Similarly, the side effects paraesthesia [67] and leukopenia [61, 68] associated with the drug vancomycin were also detected in our experiment. Moreover, Table 10 shows that the side effects somnolence [62][p. 1211], anorexia [64], and convulsion [65] associated with the drug cisplatin were found in our experiment. In addition, the side effects leukoplakia [63] and angioedema [69] associated with the drug glimperide were also found in our experiment, too.

**Table 9.** New predictions not seen in the dataset SIDER

| Vancomycin |  |  | Amlodipine |  |  |
| --- | --- | --- | --- | --- | --- |
| Side effect | Probabilities | Literatures | Side effect | Probabilities | Literatures |
| Decreased appetite | 1 |  | Diarrhoea | 1 | [57] |
| Paraesthesia | 1 | [67] | Somnolence | 1 |  |
| Dyspepsia | 1 |  | Feeling abnormal | 1 |  |
| Musculoskeletal discomfort | 1 |  | Decreased appetite | 0.999999 |  |
| Anorexia | 1 |  | Paraesthesia | 0.999999 |  |
| Hyperhidrosis | 1 |  | Dyspepsia | 0.999999 | [58, 59] |
| Diarrhoea | 1 |  | Anorexia | 0.999999 |  |
| Somnolence | 1 |  | Musculoskeletal discomfort | 0.999999 | [60] |
| Feeling abnormal | 1 |  | Hyperhidrosis | 0.999999 |  |
| Leukopenia | 0.999999 | [61, 68] | Convulsion | 0.999999 |  |

**Table 10.** New predictions not seen in the dataset SIDER, continued

| Cisplatin |  |  | Glimperide |  |  |
| --- | --- | --- | --- | --- | --- |
| Side effect | Probabilities | Literatures | Side effect | Probabilities | Literatures |
| Diarrhoea | 1 |  | Shock | 0.999999 |  |
| Somnolence | 0.999999 | [62] | Dry mouth | 0.999998 |  |
| Decreased appetite | 0.999999 |  | Leukopenia | 0.999998 | [63] |
| Feeling abnormal | 0.999999 |  | Confusional state | 0.999998 |  |
| Dyspepsia | 0.999999 |  | Oedema | 0.999998 |  |
| Paraesthesia | 0.999999 |  | Angioedema | 0.999997 | [69] |
| Musculoskeletal discomfort | 0.999999 |  | Discomfort | 0.999996 |  |
| Anorexia | 0.999999 | [64] | Agitation | 0.999996 |  |
| Hyperhidrosis | 0.999999 |  | Vision blurred | 0.999996 |  |
| Convulsion | 0.999999 | [65] | Hypertension | 0.999996 |  |

Tables 9–10 show that quite a few new side effects associated with the four drugs which were not found in the original datasets could be obtained using the GCNMLP.

## Conclusions

We have presented the GCNMLP as an improved method to predict drug side effects on the datasets SIDER, OFFSIDES, and FAERS. Since the information structure is of network type, such as protein-protein, drug-target, or drug-genome interaction [15, 66], the biomedical data is particularly suitable for the GCNMLP analysis. Except on the evaluation scale Precision, the GCNMLP outperforms traditional methods, such as matrix factorization methods, network link-prediction methods and machine learning methods implemented in this paper on the other evaluation scales. We have demonstrated novel predictions from our model, some of which can be validated in the literatures as shown in Tables 9–10. The predicting results show the capabilities of the GCNMLP, which reveals potential link information when combined with heterogenous graphs.

However, the data we chose in our study are only limited to the drug-drug similarity and drug side effect. We may extend the database by including the drug-protein or pharmacogenetics in our study to make a better personalized prediction. Moreover, we may perform the permutation test for the invariant or equivariant nature of graphs [70, 71] in our future study.

In conclusion, our study suggests that the GCNMLP might be used as a model for exploring drug side effects. We can give better personalized medicine using the GCNMLP approach.

## Data Availability

The datasets in this study were downloaded from open resources, where the drug information was obtained from DrugBank Online (\url{https://go.drugbank.com/}), and the side effects information from three Adverse Drug Event (ADE) on three databases: the Side Effect Resource (SIDER) (\url{http://sideeffects.embl.de/}), OFFSIDES \cite{OFFSIDES} (\url{http://www.pharmgkb.org/downloads.jsp}), and the United States Food and Drug Administration (FDA) Adverse Event Reporting System (FAERS) (\url{https://open.fda.gov/data/faers/}). The datasets of side effects (refer to github.com/yishingene/gcnmlp) contains four columns: `drugbank\_id' is the identification number of the database from the University of Alberta, `drugbank name' is the drug name, `umls cui from meddra' is the coded number of the Unified Medical Language System, and `side\_effect\_name' is the reported side effects.

https://github.com/timilsinamohan/sideeffects/tree/master/data

## Acknowledgments

The authors thank the anonymous referees for their kind comments and valuable suggestions. Research by the second author is supported by the Ministry of Science and Technology of Taiwan under Grants MOST 110-2634-F-005 -006 and MOST 109-2115-M-005-003-MY2.

## Author Contributions

Conceived and designed the experiments: YHC YTS CST. Performed the experiments: YHC. Analyzed the data: YHC YTS. Contributed materials/analysis tools: YHC YTS. Wrote the paper: YHC YTS CSC.

## References

1. Edwards IR & Aronson JK. Adverse drug reactions: definitions, diagnosis, and management. Lancet. 2000;356(9237):1255–9.

2. Friedman MA, Woodcock J, Lumpkin MM, Shuren JE, Hass AE, & Thompson LJ. The safety of newly approved medicines: do recent market removals mean there is a problem? Jama. 1999;281(18):1728–34.

3. Scheiber J, Chen B, Milik M, Sukuru SC, Bender A, Mikhailov D, Whitebread S, Hamon J, Azzaoui K, Urban L, Glick M, Davies JW, & Jenkins JL. Gaining insight into off-target mediated effects of drug candidates with a comprehensive systems chemical biology analysis. Journal of chemical information and modeling. 2009;49(2):308–317.

4. Bailey J, Thew M & Balls M. An analysis of the use of animal models in predicting human toxicology and drug safety. Alternatives to Laboratory Animals. 2014;42(3):181–199.

5. Gómez-Lechón MJ, Tolosa L, Conde I & Donato MT. Competency of different cell models to predict human hepatotoxic drugs. Expert Opin. Drug Metab. Toxicol. 2014;10:1553–1568.

6. Atias N & Sharan R. An algorithmic framework for predicting side effects of drugs. J. Comput. Biol. 2011;18:207–218.

7. Zhang W, Zou H, Luo L, Liu Q, Wu W & Xiao W. Predicting potential side effects of drugs by recommender methods and ensemble learning. Neurocomputing 2016;173:979–987.

8. Yamanishi Y, Pauwels E & Kotera M. Drug side-effect prediction based on the integration of chemical and biological spaces. J. Chem. Inf. Model. 2012;52:3284–3292.

9. Zhang W, Liu W, Chen Y, Wu W, Wang W & Li X. Feature-derived graph regularized matrix factorization for predicting drug side effects. Neurocomputing. 2018;287:154–162.

10. Muñoz E, Nováček V & Vandenbussche PY. Using Drug Similarities for Discovery of Possible Adverse Reactions. AMIA Annu. Symp. Proc. American Medical Informatics Association 2016. 2016;924–933.

11. Campillos M, Kuhn M, Gavin A-C, Jensen LJ & Bork P. Drug target identification using side-effect similarity. Science. 2008;321:263–266.

12. White RW, Wang S, Pant A, Harpaz R, Shukla P, Sun W, DuMouchel W & Horvitz E. Early identification of adverse drug reactions from search log data. J. Biomed. Inform. 2016;59:42–48.

13. Vilar S, Friedman C & Hripcsak G. Detection of drug–drug interactions through data mining studies using clinical sources, scientific literature and social media. Brief. Bioinform. 2017;19:863–877.

14. Vine LD, Zuccon G, Koopma n B, Sitbon L & Bruza P. Medical Semantic Similarity with a Neural Language Model. In: Proceedings of the 23rd ACM International Conference on Conference on Information and Knowledge Management. 2014 Nov 3; p. 1819–1822.

15. Mizutani S, Pauwels E, Stoven V, Goto S & Yamanishi Y. Relating drug-protein interaction network with drug side effects. Bioinformatics 2012; 28(18): i522–i528.

16. Menon AK & Elkan C. Link Prediction via matrix factorization. Machine Learning and Knowledge Discovery in Databases, Springer, Berlin and Heidelberg; Springer-Verlag.; 2011.

17. Cai J-F, Candès EJ & Shen Z. A singular value thresholding algorithm for matrix completion. SIAM J. Optim. 2010; 20:1956–1982.

18. Xu Y, Yin W, Wen Z & Zhang Y. An alternating direction algorithm for matrix completion with nonnegative factors. Front. Math. China. 2012;7:365–384.

19. Karatzoglou A, Amatriain X, Baltrunas L & Oliver N. Multiverse recommendation: n-dimensional tensor factorization for context-aware collaborative filtering. In: Proceedings of the Fourth ACM Conference on Recommender Systems, 2010. p. 79–86.

20. Grover A & Leskovec J. node2vec: Scalable feature learning for networks. In: Proceedings of the 22nd ACM SIGKDD International Conference on Knowledge Discovery and Data Mining, 2016. p. 855–864.

21. Wang H, Gu Q, Wei J, Cao Z & Liu Q. Mining drug-disease relationships as a complement to medical genetics-based drug repositioning: Where a recommendation system meets genome-wide association studies. Clin. Pharmacol. Ther. 2015; 97:451–454.

22. Nitsch D, Gonçalves JP, Ojeda F, de Moor B & Moreau Y. Candidate gene prioritization by network analysis of differential expression using machine learning approaches. BMC Bioinformatics. 2010;11:460.

23. Lovász L. Random walks on graphs: a survey, combinatorics, Paul Erdos is eighty. Bolyai Soc. Math. Stud. 1993;2:1–46.

24. Page L, Brin S, Motwani R & Winograd T. The pagerank citation ranking: bringing order to the web. Tech. Rep. Stanford InfoLab; 1999.

25. Stanfield Z, Coşkun M & Koyutürk M. Drug response prediction as a link prediction problem. Sci. Rep. 2017;7:40321.

26. Zhou J, Cui G, Hu S, Zhang Z, Yang C, Liu Z, Wang L, Li C & Sun M. Graph neural networks: A review of methods and applications. AI Open. 2020;1:57–81.

27. Hasibi R & Michoel T. A graph feature auto-encoder for the prediction of unobserved node features on biological networks. BMC Bioinform. 2021;22:525.

28. Fan W, Ma Y, Li Q, He Y, Zhao E, Tang J & Yin D. Graph neural networks for social recommendation. In: The World Wide Web Conference on - WWW ‘19. 2019 May 13; p. 417–426.

29. Wang X, He X, Cao Y, Liu M & Chua T-S. KGAT: Knowledge Graph Attention Network for Recommendation. In: Proceedings of the 25th ACM SIGKDD International Conference on Knowledge Discovery & Data Mining - KDD ‘19. 2019 Jul 25. p. 950–958.

30. Venkatesan R, Li B. Convolutional Neural Networks in Visual Computing: A Concise Guide. Boca Raton; CRC Press; 2017.

31. Zhang S, Tong H, Xu J. et al. Graph convolutional networks: a comprehensive review. Comput Soc Netw 2019; 6:11.

32. Zitnik M, Agrawal M & Leskovec J. Modeling polypharmacy side effects with graph convolutional networks. Bioinformatics. 2018; 34(13): i457–i466.

33. Muñoz E, Nováček V & Vandenbussche PY. Facilitating prediction of adverse drug reactions by using knowledge graphs and multi-label learning models. Brief. Bioinform. 2019; 20(1):190–202.

34. Mohsen A, Tripathi LP & Mizuguchi K. Deep learning prediction of adverse drug reactions in drug discovery using open TG–GATEs and FAERS databases. Front. Drug. Discov. 2021; 3.

35. Dey S, Luo H, Fokoue A, Hu J & Zhang P. Predicting adverse drug reactions through interpretable deep learning framework. BMC Bioinformatics 2018;19(21):1–13.

36. Guney E. Reproducible drug repurposing: When similarity does not suffice. Pac. Symp. Biocomput. 2017;22:132–143.

37. Wishart DS et al. DrugBank: a knowledgebase for drugs, drug actions and drug targets. Nucleic Acids Res 2008; 36(suppl 1):D901–D906.

38. Wang Y, Xiao J, Suzek TO, Zhang J, Wang J & Bryant SH. PubChem: a public information system for analyzing bioactivities of small molecules. Nucleic Acids Res. 2009;37(suppl 2):W623–W633.

39. Kuhn M, Campillos M, Letunic I, Jensen LJ & Bork P. A side effect resource to capture phenotypic effects of drugs. Mol. Syst. Biol. 2010;6(1):343.

40. Galeano D & Paccanaro A. A recommender system approach for predicting drug side effects. In: 2018 International Joint Conference on Neural Networks (IJCNN) 2018 Jul 8; p. 1–8. IEEE.

41. Nguyen T, et al. GraphDTA: predicting drug-target binding affinity with graph neural networks. Bioinformatics. 2021; 37:1140–1147.

42. Schlichtkrull M, et al. Modeling relational data with graph convolutional networks. The Semantic Web, Springer International Publishing, 2018.

43. Timilsina M, Tandan M, d’Aquin M & Yang H. Discovering links between side effects and drugs using a diffusion based method. Sci. Rep. 2019;9:10436.

44. Wishart DS, et al. DrugBank: a knowledge base for drugs, drug actions and drug targets. Nucleic Acids Res. 2008; 36:D901–D906.

45. Kuhn M, Letunic I, Jensen LJ & Bork P. The SIDER database of drugs and side effects. Nucleic Acids Res. 2016;44(D1):D1075–D1079.

46. Davis AP, King BL, Mockus S, Murphy CG, Saraceni-Richards C, Rosenstein M, Wiegers T & Mattingly CJ. The Comparative Toxicogenomics Database: update 2011 Nucleic Acids Res. 2011;39:D1067–1072.

47. Pyysalo S, Ginter F, Moen H, Salakoski T & Ananiadou S. Distributional semantics resources for biomedical text processing. In: Proceedings of LBM 2013, 2013; p. 39–44.

48. Ma H, Yang H, Lyu MR & King I. Mining social networks using heat diffusion processes for marketing candidates selection. In: Proceedings of the 17th ACM Conference on Information and Knowledge Management. 2008 Oct 26; p. 233–242.

49. Adamic LA & Adar E. Friends and neighbors on the web. Social Networks. 2003; 25(3):211–230.

50. Zhou T, Lu L & Zhang YC. Predicting missing links via local information. Eur. Phys. J. B. 2009; 71:623–630.

51. Katz L. A new status index derived from sociometric analysis. Psychometrika. 1953; 18,39–43.

52. Jeh G & Widom J. Scaling personalized web search. In: Proceedings of the 12th international conference on World Wide Web. 2003 May 20; p. 271–279.

53. Fogaras D, Rácz B, Csalogány K & Sarlaós T. Towards scaling fully personalized pagerank: Algorithms, lower bounds, and experiments. Internet Mathematics, 2005;2(3):333–358.

54. Hamilton WL, Ying R & Leskovec J. Inductive representation learning on large graphs. Advances in neural information processing systems. 2017;30.

55. Kingma DP & Ba J. Adam: a method for stochastic optimization. [cs.LG]:1412.6980. 2014 Dec 22.

56. Defferrard M, Bresson X & Vandergheynst P. Convolutional neural networks on graphs with fast localized spectral filtering. In Proceedings of the 30th International Conference on Neural Information Processing Systems, 2016; 29:3844–3852.

57. Russell RP. Side effects of calcium channel blockers. Hypertension. 1988; 11:II42.

58. Hosie J, et al. Comparison of early side effects with amlodipine and nifedipine retard in hypertension. Cardiology 80 Suppl. 1992;1:54–59.

59. Devasahayam J, Pillai U & Uppaluri C. Acute severe intestinal obstruction secondary to amlodipine toxicity. QJM. 2012;105:467–469.

60. Phillips BB & Muller BA. Severe neuromuscular complications possibly associated with amlodipine. Annals of Pharmacotherapy. 1998;32(11):1165–1167.

61. Kemeç Z, Kahya Y, Atay A, Demir M & Gürel A. Vancomycin dependent pancytopenia-a rare side effect: a case report. International Journal of Medical Reviews and Case Reports. 2019;3:1.

62. Biller J & Ferro JM. Neurologic Aspects of Systemic Disease, Part III. Handbook of Clinical Neurology. 2014.

63. Dwivedi K, Saraswat N & Bisht M. Protons confirmation of glimepiride drug using correlation spectroscopy a unique tool of nuclear magnetic resonance spectroscopy. Math. Sci. Res. J. 2013; 3:1–9.

64. Saito H. Autonomic dysreflexia in a case of radiation myelopathy and cisplatin-induced polyneuropathy. Spinal Cord Ser Cases 2020;6(1):1–5.

65. Ishihara M, Matsutani N, Ota S & Seki N. A case of posterior reversible encephalopathy syndrome induced by cisplatin/cpemetrexed chemotherapy for lung cancer. Case Rep. Oncol. 2017;10:235–238.

66. Turanli B et al. A network-based Cancer drug discovery: from integrated multi-omics approaches to precision medicine. Curr. Pharm. Des. 2018;24:3778–3790.

67. Cohen LG, Souney PF & Taylor SJ. Paresthesia and back pain in a patient receiving vancomycin during hemodialysis. Drug Intell. Clin. Pharm. 1988;22:784–785.

68. Hook EW 3rd & Johnson WD Jr. Vancomycin therapy of bacterial endocarditis. Am. J. Med. 1978;65:411–415.

69. Handelsman Y, et al. A randomized, double-blind, non-inferiority trial evaluating the efficacy and safety of omarigliptin, a once-weekly DPP-4 inhibitor, or glimepiride in patients with type 2 diabetes inadequately controlled on metformin monotherapy. Curr. Med. Res. Opin. 2017;33:1861–1868.

70. Niu C et al. Permutation invariant graph generation via score-based generative modeling. In: International Conference on Artificial Intelligence and Statistics 2020 Jun 3; p. 4474–4484. PMLR.

71. Keriven N & Peyrè G. Universal invariant and equivariant graph neural networks. Advances in Neural Information Processing Systems. 2019;32.

